# Supplementary motor area microstructure as measured by diffusion imaging defines the extent of gait impairment in Parkinson’s disease

**DOI:** 10.1101/2025.02.11.25321668

**Authors:** Paweł P. Wróbel, Annika Peter, Maja Kirsten, Alessandro Gulberti, Maxim Bester, Einar Goebell, Bastian Cheng, Yogesh Rathi, Ofer Pasternak, Tim Magnus, Götz Thomalla, Fanny Quandt, Robert Schulz, Focko L. Higgen, Monika Pötter-Nerger

**Author notes:** **Corresponding authors:** Paweł Wróbel MD; University Medical Center Hamburg-Eppendorf; Martinistr. 52, 20246 Hamburg, Germany;, Monika Pötter-Nerger MD; University Medical Center Hamburg-Eppendorf; Martinistr. 52, 20246 Hamburg, Germany. These authors contributed equally to this work (Focko L. Higgen, Monika Pötter-Nerger).

## Abstract

**Background:** Gait disorders are therapeutically challenging symptoms in Parkinson’s disease. However, the pathophysiological mechanisms and the interaction between cerebral structure and clinical burden are not yet fully understood. Objectives: Given that cortical microstructure is a crucial determinant of symptom burden in neurodegenerative diseases, we hypothesized that diffusion-imaging-based microstructural markers in the supplementary motor area, one cortical key hub in the gait network, are associated with gait performance. Methods: In this retrospective study, 29 patients (mean age 64.1 ± 6.9 years, 11 (37.9%) female, mean disease duration 10.9 ± 4.5 years) were included. Diffusion-tensor imaging measures and cortical thickness (fractional anisotropy, radial and axial diffusivity) in the supplementary motor area as defined by the Human Motor Area Template were correlated with quantitative gait performance parameters. Results: Lower anisotropy in the right supplementary motor cortex was significantly associated with better performance across various bilateral gait parameters during walking at natural speed, at maximal velocity and particularly during dual-tasking. There were no significant associations between cortical thickness and gait performance. Conclusions: Given the inverse relationship between anisotropy and cortical tissue complexity, the observed lower anisotropy in the right supplementary motor area of patients with better gait performance supports the assumption of the modulatory role of this cortical area in gait control suggesting a potential therapeutic target e.g. stimulation, in Parkinson’s disease patients with gait disorders.

## Introduction

The impairment of gait is a fundamental aspect of symptom burden that affects the quality of life in Parkinson’s Disease (PD) patients^1^ and the pathophysiology of gait disorders remains not entirely understood^2^. Within the neuronal network of gait control, the supplementary motor area (SMA) has been identified as a key hub for producing, modulating and transmitting motor programs necessary for purposeful gait^3^, especially in conditions with increased cognitive load^4^. Anatomically, the SMA is strongly connected to the pedunculopontine nucleus (PPN) in the mesencephalic locomotor region through direct and indirect projections via the basal ganglia^2,5^. The functional link between the SMA and the mesencephalic locomotor region has been demonstrated through the functional coupling of bilateral SMA in patients with gait disorders^6^ and in white matter disconnection syndromes in animals and humans^7-9^. SMA activity was also associated with the initiation and sequencing of voluntary movements^10,11^.

Positron emission tomography studies showed a positive relationship between SMA metabolism and gait performance in healthy adults and those with gait disturbances^12,13^, particularly for step length^14,15^. In fMRI studies BOLD signal from SMA was related to lower step length^16,17^ and SMA pathology has been implicated in the manifestation of gait impairment, as seen in studies involving ischemic lesioning^18^ or corticectomy^19^. Taken together, the SMA appears to serve as an important cortical interface in cognitive gait control and alterations in the SMA structure could be a relevant pathophysiological cursor of gait disorders.

However, imaging studies on cortical atrophy topology and gait impairment in PD report varying results, without clear patterns^20-22^. Of note, even a large meta-analysis of 1843 patients and 1172 healthy controls found no differences in cortical thickness (CT) between PD patients and controls across the cortex, stating CT to be an unsuitable to grasp PD pathology^23^.

The heterogeneous results indicate that other approaches, such as microstructural surrogates of complexity, are necessary to identify cortical pathology. Diffusion-tensor imaging (DTI) offers more sensitive measures that may grasp microstructural correlates, as indicated by the correspondence of the diffusion and histology tensors in comparative studies^24,25^. For instance, axial diffusivity (AD) represents diffusion along the tensor and therefore neuronal columns, and reflects the density of apical dendrites^26^. Radial diffusivity (RD) occurs orthogonally to neuronal columns and correlates positively with the number of neuronal structures and thus the degree of dendritic arborization^25^. Fractional anisotropy (FA) is a scalar value expressing the anisotropy of water diffusion, which is influenced positively by AD and negatively by RD. To date, an increase in cortical FA has been shown in aging^27^, cortical pruning^28^, and after concussion^29^, while lower FA was reported during maturation-associated dendritic spreading^30,31^.

Considering the literature on electrophysiologic and metabolic SMA activation being related to gait performance, along inconsistent reports on macrostructure, this retrospective work assesses the structure-function relationship of SMA and gait based on the hypothesis that microstructural features of the SMA, as detected by DTI, will be correlated with objective gait characteristics in PD patients. As PD is associated with compromised gait parameters^32^, further confirmation of SMA as key structure for gait impairment would be of great therapeutic relevance, for instance for local stimulation, given the disability-driving gait impairment and its frequent resistance to treatment.

## Methods

### Participants & clinical data

A total of 32 PD patients who underwent standardized gait evaluation and had available MRI datasets, collected for perioperative tractography between July 2020 and October 2022, were initially identified for the retrospective analysis of diffusion-weighted imaging (DWI) data. The imaging and clinical data acquisition was performed within 8 days, except for two patients for whom the data was acquired with an interval of 10 (patient 21) and 6 (patient 26) months due to regulations SARS-CoV-2 pandemic. Following manual quality assessment, three datasets were excluded due to insufficient structural MRI quality, which hindered the identification of the gray/white matter boundary.

The final cohort comprised 29 patients (mean age 64.1 ± 6.93 years, 37.93% female, mean disease duration 10.93 ± 4.50 years). A single patient’s dual-task data was not available, thus resulting in 28 datasets in the dual-task analyses. Detailed demographic and clinical information is provided in Table 1.

**Table 1.**
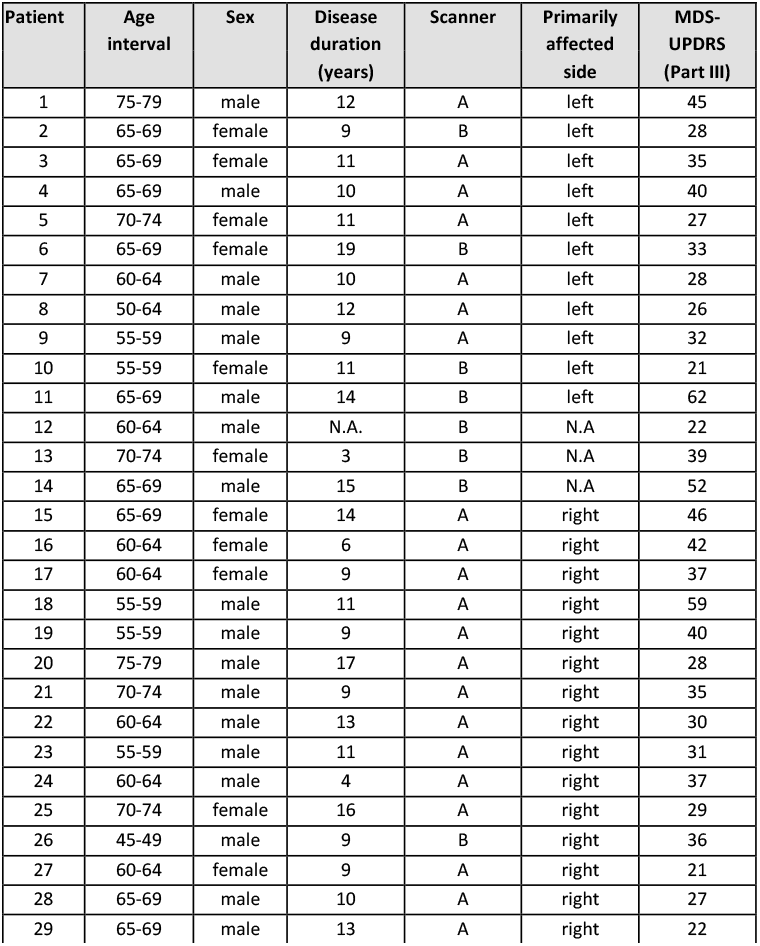
Demographic and clinical data of the patients. Age range (years): 47 – 75 (mean: 64.1, median: 65.0, standard deviation (SD): 6.93. Age ranges are given for deidentification purposes. Gender ratio (♂:♀): 62.07%:37.93%. UPDRS Part III OFF-phase: 21 - 62 (mean: 34.83, median: 43.00, SD: 10.54) – rated by the same physician. N.A. – not available. Scanner A: 3 Tesla Siemens Prisma Scanner (Siemens Healthineers, Erlangen, Germany). Scanner B: 3 Tesla Siemens Skyra Scanner (Siemens Healthineers, Erlangen, Germany).

| Patient | Age interval | Sex | Disease duration (years) | Scanner | Primarily affected side | MDS-UPDRS (Part III) |
| --- | --- | --- | --- | --- | --- | --- |
| 1 | 75-79 | male | 12 | A | left | 45 |
| 2 | 65-69 | female | 9 | B | left | 28 |
| 3 | 65-69 | female | 11 | A | left | 35 |
| 4 | 65-69 | male | 10 | A | left | 40 |
| 5 | 70-74 | female | 11 | A | left | 27 |
| 6 | 65-69 | female | 19 | B | left | 33 |
| 7 | 60-64 | male | 10 | A | left | 28 |
| 8 | 50-64 | male | 12 | A | left | 26 |
| 9 | 55-59 | male | 9 | A | left | 32 |
| 10 | 55-59 | female | 11 | B | left | 21 |
| 11 | 65-69 | male | 14 | B | left | 62 |
| 12 | 60-64 | male | N.A. | B | N.A | 22 |
| 13 | 70-74 | female | 3 | B | N.A | 39 |
| 14 | 65-69 | male | 15 | B | N.A | 52 |
| 15 | 65-69 | female | 14 | A | right | 46 |
| 16 | 60-64 | female | 6 | A | right | 42 |
| 17 | 60-64 | female | 9 | A | right | 37 |
| 18 | 55-59 | male | 11 | A | right | 59 |
| 19 | 55-59 | male | 9 | A | right | 40 |
| 20 | 75-79 | male | 17 | A | right | 28 |
| 21 | 70-74 | male | 9 | A | right | 35 |
| 22 | 60-64 | male | 13 | A | right | 30 |
| 23 | 55-59 | male | 11 | A | right | 31 |
| 24 | 60-64 | male | 4 | A | right | 37 |
| 25 | 70-74 | female | 16 | A | right | 29 |
| 26 | 45-49 | male | 9 | B | right | 36 |
| 27 | 60-64 | female | 9 | A | right | 21 |
| 28 | 65-69 | male | 10 | A | right | 27 |
| 29 | 65-69 | male | 13 | A | right | 22 |

### Gait parameters

Gait parameters were measured using a GAITRite® electronic walkway system (CIR Systems Inc., Franklin, New Jersey, USA). The GAITRite® system consists of a walkway with overall dimensions of 90cm×7m×3.2mm. Each patient was tested in an off situation, i.e. the dopaminergic and agonistic treatment was paused 24 hours in advance and completed three different gait tasks in a randomized order including 1) preferred self-paced gait (normal), 2) maximum speed walking without running (fast) and 3) dual-task walking while counting backward in increments of 7 (dual-task). Every task was performed three times and averaged data were used for the analyses. The analyses included the step count (absolute), step length (cm), velocity (cm/sec), cadence (steps/min) and the percentage of the stance of each leg within the gait cycle time. The data were corrected for the leg length. Detailed data are summarized in Supplementary Tables 2-4.

### Cerebral Imaging Data – Acquisition and Processing

Magnetic resonance imaging (MRI) was conducted using either a 3 Tesla Siemens Prisma Scanner (Siemens Healthineers, Erlangen, Germany) [Scanner A] or a 3 Tesla Siemens Skyra Scanner [Scanner B], both equipped with a 32-channel head coil. 3D T1-weighted imaging datasets were acquired with a magnetization-prepared, rapid acquisition gradient-echo sequence (MPRAGE) (Scanner A and B: echo time [TE]: 2.46ms, repetition time [TR]: 1900ms, 256 slices with field of view (FOV): 256×160mm, slice thickness 1mm and an in-plane resolution of 0.94×0.94mm). Diffusion MRI data were acquired with one b_0_ image and 64 non-collinear gradient directions with a b-value of 1500s/mm^2^ in a 128×104mm FOV (Scanner A: 75 slices, TE: 82ms, TR: 10700ms and voxel size of 2×2×2mm; Scanner B: 70 slices, TE: 82ms, TR: 10900ms and voxel size of 2×2×2mm).

T1-weighted structural data were segmented and parcellated using the FreeSurfer-based *recon-all* tool (version 6.0.1) to facilitate the identification of gray matter^33^. The segmentation data were then registered to the individual pre-processed (DWI) data using advanced Normalization Tools (ANTs – version 2.3.4) (Avants *et al*., 2011). The mean CT of both SMA was calculated after registering the Human Motor Atlas Template (HMAT)^34^ data to individual segmentations using FreeSurfer’s *vol2surf* and subsequently collected via *mri_segstats*.

Eddy current correction was applied for the DWI data, followed by a T1 image-based EPI distortion correction and brain extraction using MRtrix3 (3.0.2) and FSL (6.0.1)^35,36^. Free water (FW) correction, to account for the vicinity of cerebrospinal fluid, was conducted according to previously established methods ^37^ by fitting a bi-tensor model with pre-set FW diffusivity using MATLAB (Version R2020a, The Mathworks, Natick, MA, USA)^38^. The FSL-based non-linear transformation was applied to SMA labels (from Montreal Neurological Institute (MNI) space to individual DTI space) using *flirt* and *fnirt* tools, including nearest neighbor interpolation. The labels were multiplied with a cortex mask to exclude voxels from non-grey matter tissue that might have been produced during the transformation. Finally, for each SMA averaged FA, axial diffusivity (AD) and radial diffusivity (RD) values were derived from FW-corrected tensor eigenvalues using a custom-written MATLAB script (ran on Version R2020a, The Mathworks, Natick, MA, USA).

### Statistical analysis

Statistical analyses were performed using R software^39^, version 4.0.2. Linear regression models were employed to examine the relationship between gait parameters, namely velocity, cadence, step count, step length, and stance time as a percentage of the cycle time (STp), as the independent and imaging measures of SMA as dependent variables of interest. AGE and despite for the adjustment of the data for the leg length also SEX were treated as nuisance variables to adjust for potential confounding effects. For DTI measures, the models were further adjusted for CT in the corresponding SMA, to account for the potential partial volume effect of white matter, which could potentially lead to artificially increased FA in a thinner cortex. Further factors were assessed in post-hoc analyses as stated in the subsequent section. Separate models were calculated for the left and right SMA, as well as for the left and right leg for step length and the STp, resulting in a total of 42 models for each imaging measure. Results are presented after correction for multiple comparisons using the False-Discovery-Rate (FDR) with an assumed significance level at *P*_*FDR*_<0.05. As one could argue that the findings could hypothetically be attributable to other factors, further sensitivity analyses for the right SMA were conducted to assess the models’ robustness. Given potential influence of the highly-skewed factor scanner on behavioral and diffusion data the models were recalculated with an interaction term between diffusion and scanner variables. Further, a relevant microstructure-function relationship with gait could also coexist in the leg-area of the primary motor cortex. To this end, the models were recalculated for the gait data imaging data from the cortical region comprising the primary motor leg area, i.e. G_and_S_paracentral label from the Destrieux atlas. Even though the models were adjusted for sex, gait characteristics remain highly sex-specific. For instance, if somehow women were younger, or healthier, it could explain the correlations. Therefore the analyses were repeated for sex-based sub-groups. The primarily affected side was not included in the original model as the missing data would lead to underpowered, skewed and overfitted models. A multivariable model for all factors appeared also unsuitable for the same reasons. Lastly, the original models were tested by leave-one-out analyses (LOOA), to assess if the significance levels were not driven by single data sets.

### Data availability

The data used in the statistical analysis is available in the supplementary material.

## Results

The gait characteristics in PD patients showed a mean step length (left: 50.32cm ±11.86, right: 52.61cm ±12.20), mean step count (average: 11.10 ±3.23), velocity (93.21 cm/s ±23.96), changed cadence (109.88s^-1^ ±12.36), and prolonged stance time percentage of the step cycle (left: 66.24% ±3.22, right: 66.90% ±3.22) during normal gait. These deficits were exacerbated under gait conditions with increased cognitive load (Suppl. Tables 2-4).

MRI data for the left and right SMA indicated an average FA of 0.151 and 0.147 and an average CT of 2.44mm and 2.45mm respectively. The main, recurrent finding was a significant relationship between FA in the right SMA and bilateral objective gait parameters as step count, step length, velocity, cadence and percentage of the stance of each leg within the gait cycle after the FDR correction (Figures 1 and 2). No significant associations were found between FA in the left SMA and primary leg motor cortex and gait parameters after correction. MR based CT analysis did not yield any significant results with clinical gait parameters. Of note, general motor symptoms reflected by the MDS-UPDRS Part III were not linked to FA of the right SMA. In the post-hoc analysis FA of the primary leg motor cortex and objective gait parameters did not reveal any significant relationships or trends after correction. A supplementary analysis revealed no significant effects of the skewed factor SCANNER, even at the uncorrected level. However, interactions with FA were detected in the models for gait velocity under dual-task conditions and for the stance percentage of the left leg (regular and dual-task). In the post-hoc analysis on AD and RD, there were no statistically significant findings after the FDR correction.

**Figure 1.**
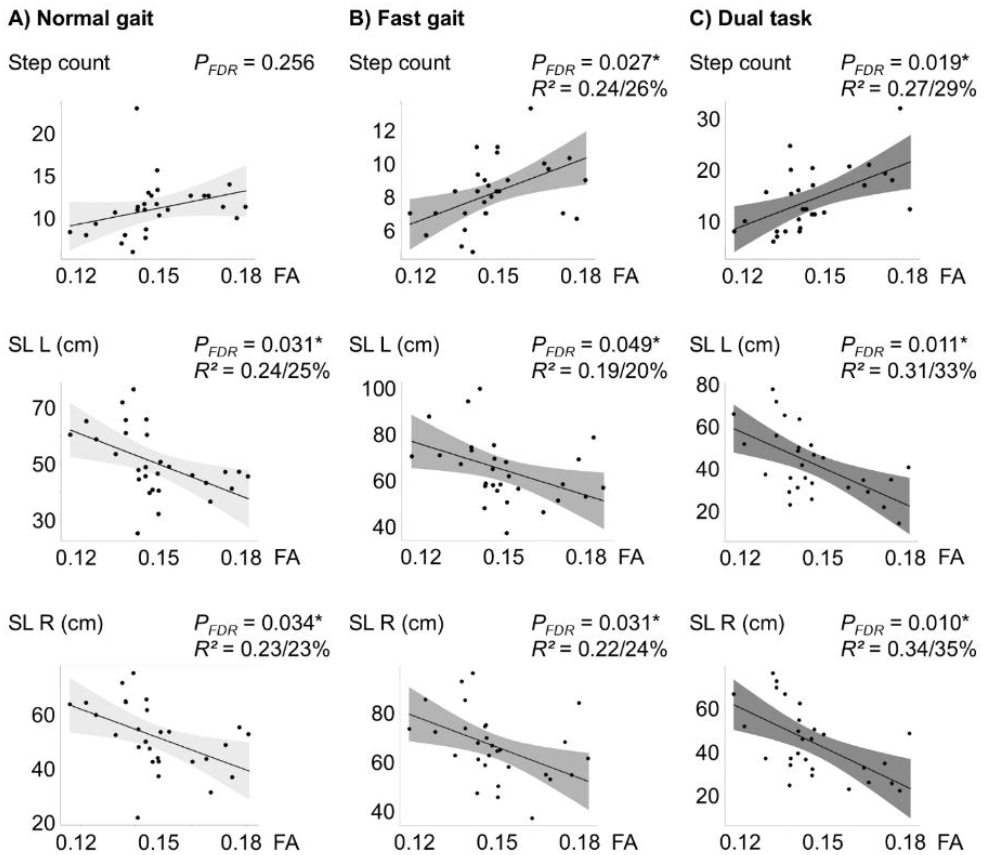
Relationship of right Supplementary Motor Area (SMA) anisotropy with step count and step length (SL). Fractional anisotropy (FA) values are plotted against step count and step length (SL) for the right (SL R) and left (SL L) leg under three conditions: A) normal gait, B) fast gait, and C) dual-task. Lower FA values, are associated with fewer and longer steps. Raw data points are presented along the model fit. *P*_*FDR*_ - P-value corrected for False-Discovery-Rate (FDR), *R*^2^ - the partial *R*^2^ value for FA from the model - the amount of unexplained variance reduced by the FA value is appended as a percentage.

**Figure 2.**
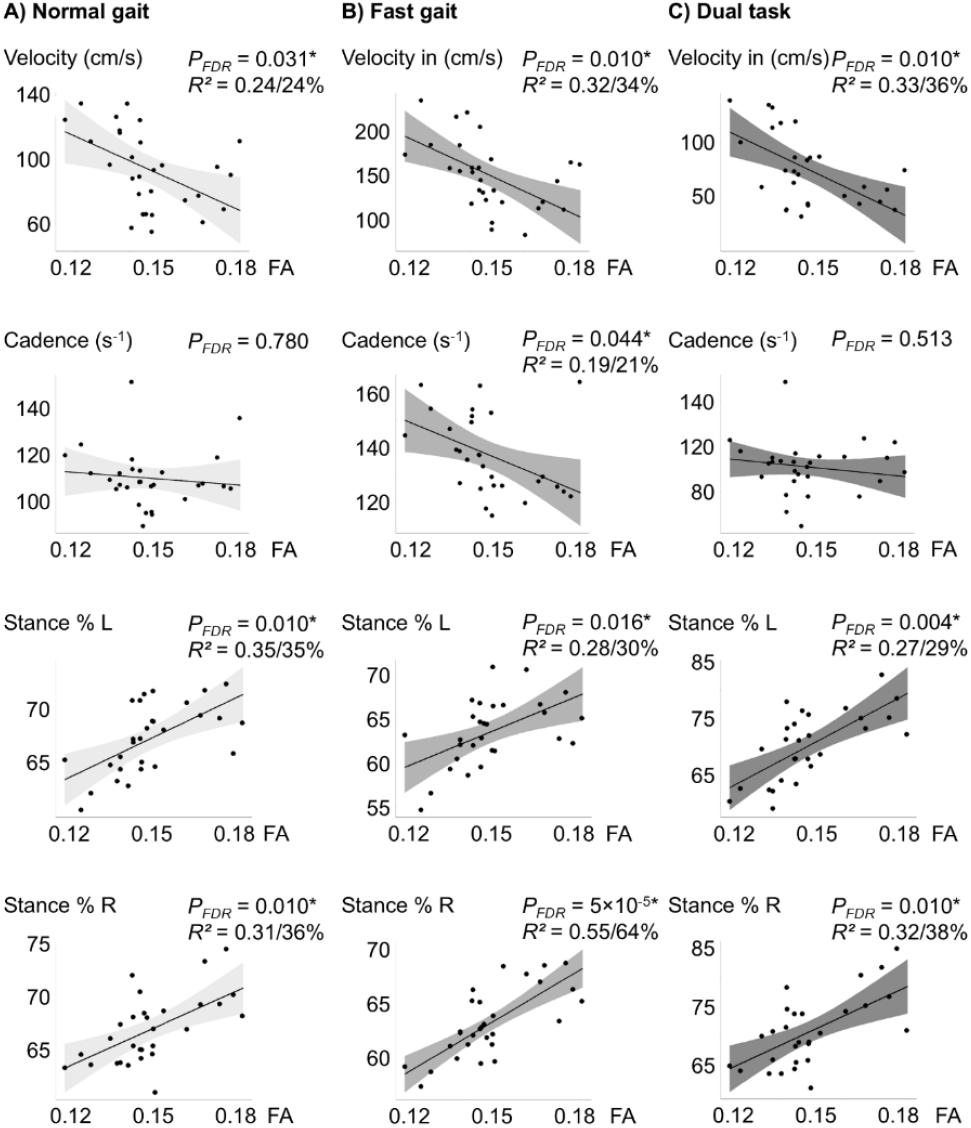
Relationship of right Supplementary Motor Area (SMA) anisotropy with velocity, cadence and stance. Fractional anisotropy (FA) values were plotted against gait velocity, cadence, and the percentage of stance time during the step cycle for each leg under three conditions: A) regular gait, B) fast gait and C) dual-task. Lower FA values, indicative of greater microstructural complexity, are associated with faster gait velocity, higher cadence and shorter stance times. Raw data points are presented along the model fit. *P*_*FDR*_ - P-value corrected for False-Discovery-Rate (FDR), *R*^2^ - the partial *R*^2^ value for FA from the model - the amount of unexplained variance reduced by the FA value is appended as a percentage.

### FA of right SMA and its relation to gait

Visualized plots including raw data are shown in Figures 1 and 2. Fewer and longer steps of both legs during fast and dual-task gait were associated with lower FA. During normal gait this finding was also observed for both legs’ step length but not the total step count. The reduction of unexplained variance with the right SMA FA values in the models ranged between 20-35%. Greater gait velocity was significantly correlated with lower FA in the right SMA across normal, fast and dual-task gait, with FA accounting for 24% of the unexplained variance in normal, 34% in fast walking and 36% in the dual-task condition. Cadence was significantly explained by FA in the right SMA only during fast gait, accounting for a 21% reduction in unexplained variance. Lastly, patients with lower stance time as percentage of the gait cycle showed lower right SMA FA. This was observed for both legs and in all three testing conditions with reduced variance between 29-64%. The results were reproduced in the LOOA approach except for the step length of the left leg and cadence in fast gait conditions, which showed a trend.

### Sex-specificity

With the aforementioned suspected influence of gender on gait characteristics, we performed a sensitivity analysis in sex-based subgroups. The testing in sex-based subgroups replicated significant findings in 15 tests in 18 males, for most gait characteristics. The data from the smaller group of 11 female participants reproduced significant findings in the fast condition for every gait parameter, except the percentage of the stance of the right leg with nearly significance. Due to the smaller sample sizes, the results were not corrected.

## Discussion

This retrospective evaluation of clinical and imaging data aimed to investigate if operationalized objective gait measures associated with gait disturbance in PD patients correlate with surrogates of microstructure in the SMA. The data revealed significantly lower FA in the right SMA of patients with longer steps, higher gait velocity and a reduced fraction of stance during the step cycle.

### Gait impairment and SMA microstructure

A significant relationship between gait performance and SMA FA appears plausible considering the role of SMA activity in gait control^5,14,15^. The data suggest that microstructural but not macroscopic measures of SMA, i.e. FA but not CT, are associated with function within the neuronal gait control network. Lower FA in the right SMA of PD patients was correlated with better gait performance, which could be interpreted in different ways. As FA is reported to decrease in dendritic spreading during cortical maturation^30,31^ and increase in aging^27^ and during cortical pruning^28^, lower FA could be discussed as a correlate for a more complex SMA in better-performing patients. A more complex histoarchitecture might facilitate more effective and efficient processing within the SMA microcircuits, leading to the aforementioned relationship between SMA activity and gait performance. Efficient local processing at the SMA could potentially modulate gait-related macrocircuits more effectively, resulting in a smoother gait (for a conceptual representation, see Figure 3).

**Figure 3.**
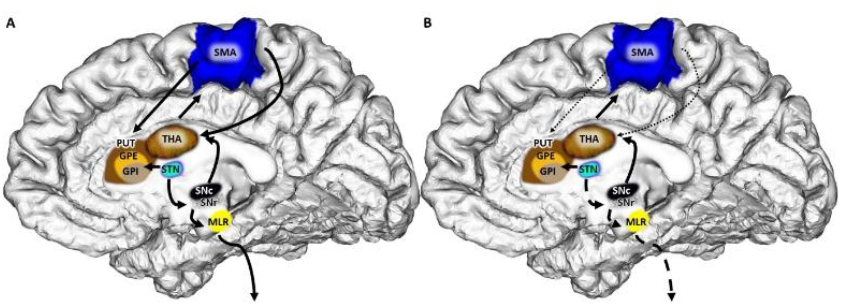
Influence of the right Supplementary Motor Area (SMA) on the pathophysiology of gait impairment. The lower fractional anisotropy (FA) of the right-hemispheric SMA in individuals with better gait performance could be interpreted as a sign of greater cortical complexity that aids in controlling gait. Compared to physiologic state (A), Parkinson’s Disease patients (B) with compromised SMA microstructure, display deteriorated control over the Striatum and Subthalamic nucleus, potentially facilitating gait impairment. THA: Thalamus, PUT: Putamen, GPE/I: Globus pallidus externus/internus. STN: Subthalamic nucleus. SNc/r: Substantia nigra, pars compacta/ reticularis, MLR: mesencephalic locomotor region. Not shown: the caudate nucleus.

Of note, the negative finding for DTI measures in the primary leg motor area implies that the structure-function relationship for gait modulation might be specifically associated with SMA. This could correspond to the indirect pathway of the gait model which is proposed to be involved in the control of gait adaptation to environmental changing conditions, between secondary motor regions and subcortical grey matter, whose dysfunction was proposed to contribute to gait pathology^40^.

The association between cortical complexity and integrative specialization has already been supported by histological studies dating back to Brodmann, which showed that integrative secondary heteromodal cortices are characterized by a greater amount of interconnected structures compared to non-integrative primary unimodal areas^41,42^. Our earlier work on DTI and cortical specialization supports this supposition^43^. Of note, as also pointed out in our earlier work, cortical complexity does not necessarily equal a higher quantity of tissue. In neurodegenerative disease, a less prominent directionality within the cortex could be also considered a degeneration^44^. The negative findings for AD and RD in this study could be linked to either a small sample size or heterogeneous processes detectable only by FA as a ratio measure. Taken together, one could suspect that the lower FA in better-performing patients could be interpreted as a sign of structural reserve^45^, leading to better performance given the preserved capacity. Still, these assumptions remain speculative and future studies will be needed to replicate the findings from this explorative work.

### Modulatory role of the right SMA in gait control

Numerically, the models for fast and dual-task gait characteristics and SMA microstructure frequently showed greater effect sizes and the most significant findings or trends. If the lower cortical anisotropy is linked to better performance, especially during increased cognitive load, it could be discussed as a correlate of a cognitive load-dependence of the gait modulation by SMA. Behavioral studies suggest that gait worsens in the presence of visual obstacles^4^. Existing stimulation studies showed that temporary impairment of SMA influences the design of complex motor tasks^46^. In the clinical setting, the suspected modulatory role of the SMA on gait in PD has already been postulated in combined PET/EEG studies^47^, especially in the right hemisphere. On a speculative note, it could be argued that microstructural complexity, as reflected in preserved interneuronal connections^30^, facilitates greater integrative capabilities or compensation which was reproduced with numerically greater values in cognitive load trials. A limitation might be the collinearity among the gait characteristics^48^, which could explain the observations within each single trial, however, we interpret the reproduction of similar findings within the three different, randomized gait tasks as a measure of reliability of the SMA-gait dependence at a qualitative level.

### Lateralization of SMA to the right hemisphere

Strikingly, the positive gait correlations were limited to the right SMA, suggesting potential cerebral hemispheric specialization. While EEG studies indicate SMA involvement in externally paced bilateral foot movements^49^, their poor spatial resolution limits the analysis of both regions’ contributions. Functional MRI studies provide inconsistent results, with some showing greater activation in the right^17,50^ and others in the left SMA^16^ during gait-related imagery tasks. The small sample sizes and the fact that the tasks involved only mental imagery could be confounding factors in these studies. Early photon emission tomography studies, testing actual walking, found alterations in SMA metabolism on the right side (see Fig 2. by Fukuyama *et al*.)^12^. When comparing the findings to macrostructural data, despite the lack of CT differences in PD patients reported in a meta-analysis, CT appears to decay particularly in the right but not left SMA^23^. The right-hemispheric finding in this study supports the CT data. Given the negative CT but positive DTI finding in this cohort, one could speculate that cortical DTI may be a more sensitive measure for detecting cortical pathology, as patients in this work, who were candidates for stimulation surgery were not severely affected.

### Clinical implications

Reflecting on the clinical implications for PD patients, the association between lower cortical FA which could indicate a more complex cortex and better gait performance may be interpreted as an indicator of structural reserve^45^.

From a clinical perspective, this link underscores the value of early physiotherapy as a preventive measure. Since a more complex SMA is associated with preserved performance despite for example impoverished subcortical afferents, it represents a promising target region for interventions, such as cortical stimulation techniques focused on the right SMA. The relevance of SMA as a target has already been described in literature on magnetic transcranial stimulation^51^. Given the limited responsiveness of gait impairments to pharmacologic treatment, such alternative therapeutic approaches remain important. Alternatively, one could optimize deep brain stimulation approaches for gait impairments by steering the volume of tissue activated to fiber bundles, particularly to the right SMA^52^. Further, the potential of DTI as a more suitable tool in detecting cortical disease implies diagnostic potential at earlier stages.

### Limitations

With a rather novel approach to measure surrogate parameters for microstructure, this study has several limitations. First, regarding the design, the retrospective analysis was performed without healthy controls, which will need addressing in future studies, given the lack of existing literature. Second, the study had a relatively small sample size. Therefore, we employed a conservative approach, including the LOOA and correction for multiple comparisons. Third, in light of the lateralized findings, information on the patient’s handedness would have been useful in assessing the right-hemispheric results. However, given the high percentage of right-handedness in the population, single datasets of left-handed patients are unlikely to have altered the results, particularly in light of stable LOOA results. Fourth, the data were acquired using two different scanners, which could have influenced the acquired data. Still, the sensitivity analysis addressed the issue and showed stable results. Fifth, the strength of diffusion weighting, as measured by b-values, could influence the results of this study^53^. Further research will be necessary to assess the most suitable imaging parameters for neurodegenerative diseases. Lastly, the diffusion measures can be influenced by partial volume effects from white matter. The models were therefore adjusted for cortical thickness, accounting for increased white matter signal in the thinner cortex.

## Conclusion

This work is the first to link surrogates of microstructural properties of the right SMA to gait impairment in PD patients, particularly in light of pre-existing evidence on SMA’s role in motor function planning, indicating a structure-function relationship. In contrast to the primary motor cortex with negative findings, microstructural alterations in the right SMA may explain bilateral gait control and its impairment in PD. These findings could serve as a base for new therapeutic approaches, such as right-sided cortical stimulation, to target symptoms of gait impairment.

## Data Availability

The original data are clinical data and thus not available to public. Remaining anonymized data was provided in the supplementary material.

## Acknowledgments

We thank Marlies Schütte for the assistance in the acquisition of the gait data.

