## Supplementary material for "Supplementary motor area microstructure as measured by diffusion imaging defines the extent of gait impairment in Parkinson’s disease"

* - corresponding authors

| Patient | Age | FA SMA | | CT SMA | |
|---|---|---|---|---|---|
| | | left | right | left | right |
| 1 | 75-79 | 0.1642862 | 0.14601372 | 2.610 | 2.566 |
| 2 | 65-69 | 0.17904964 | 0.14494878 | 2.167 | 2.354 |
| 3 | 65-69 | 0.13495703 | 0.12555593 | 2.480 | 2.568 |
| 4 | 65-69 | 0.1458998 | 0.12199413 | 3.034 | 2.611 |
| 5 | 70-74 | 0.14235042 | 0.14102942 | 2.016 | 2.143 |
| 6 | 65-69 | 0.15084422 | 0.16547967 | 1.892 | 2.05 |
| 7 | 60-64 | 0.14323078 | 0.14344405 | 1.806 | 1.962 |
| 8 | 50-64 | 0.14820392 | 0.1406603 | 2.318 | 2.305 |
| 9 | 55-59 | 0.13146909 | 0.15202564 | 2.453 | 2.499 |
| 10 | 55-59 | 0.14159164 | 0.14089973 | 2.880 | 2.732 |
| 11 | 65-69 | 0.16623066 | 0.18066512 | 2.221 | 2.11 |
| 12 | 60-64 | 0.1722203 | 0.17741975 | 2.366 | 2.081 |
| 13 | 70-74 | 0.14481969 | 0.1482711 | 2.684 | 2.511 |
| 14 | 65-69 | 0.15718997 | 0.16704188 | 2.639 | 2.829 |
| 15 | 65-69 | 0.14791669 | 0.14890814 | 2.574 | 2.376 |
| 16 | 60-64 | 0.14713605 | 0.14402974 | 2.709 | 2.8 |
| 17 | 60-64 | 0.14686765 | 0.14793883 | 2.551 | 2.337 |
| 18 | 55-59 | 0.1742654 | 0.11611313 | 2.327 | 2.534 |
| 19 | 55-59 | 0.15911999 | 0.13631441 | 2.628 | 2.711 |
| 20 | 75-79 | 0.14547843 | 0.14815956 | 2.510 | 2.635 |
| 21 | 70-74 | 0.16144833 | 0.17240795 | 2.657 | 2.278 |
| 22 | 60-64 | 0.16319863 | 0.13911486 | 2.192 | 2.462 |
| 23 | 55-59 | 0.13933717 | 0.13627936 | 2.275 | 2.406 |
| 24 | 60-64 | 0.15607941 | 0.17478669 | 2.433 | 2.568 |
| 25 | 70-74 | 0.1394691 | 0.13266487 | 2.351 | 2.111 |
| 26 | 45-49 | 0.1479108 | 0.16047555 | 2.763 | 3.025 |
| 27 | 60-64 | 0.1385086 | 0.14355531 | 2.438 | 2.541 |
| 28 | 65-69 | 0.13715492 | 0.1350731 | 2.665 | 2.584 |
| 29 | 65-69 | 0.1491434 | 0.14382502 | 2.147 | 2.224 |

**Supplementary Table 1 | Demographic and imaging data of the patients**.

Summary of the imaging data for each patient. Age range (years): 47 – 75. Age intervals are given for deidentification purposes. FA: fractional anisotropy, SMA: supplementary motor area. CT: cortical thickness.

| Patient | Step length - left | | | Step length - right | | | Step count | | |
|---|---|---|---|---|---|---|---|---|---|
| | N | F | D | N | F | D | N | F | D |
| 1 | 40.92 | 58.13 | 35.75 | 42.37 | 66.50 | 36.36 | 12.67 | 8 | 12.33 |
| 2 | 39.79 | 55.46 | 41.83 | 47.30 | 62.49 | 45.86 | 13 | 8.67 | 12.33 |
| 3 | 58.76 | 70.84 | 51.79 | 59.76 | 72.03 | 51.66 | 9.33 | 7 | 10 |
| 4 | 65.26 | 87.59 | 66.03 | 64.24 | 85.17 | 66.42 | 8 | 5.67 | 8 |
| 5 | 44.61 | 58.52 | 35.74 | 47.84 | 60.91 | 36.88 | 11.33 | 9.33 | 15.33 |
| 6 | 43.32 | 51.18 | 34.71 | 43.49 | 54.78 | 32.64 | 12.67 | 10 | 17 |
| 7 | 45.72 | 64.85 | 31.14 | 49.77 | 74.23 | 39.19 | 11.67 | 7.67 | 16 |
| 8 | 25.44 | 47.87 | 29.13 | 21.66 | 47.16 | 24.53 | 23 | 11 | 24.67 |
| 9 | 49.04 | 56.33 | 45.25 | 53.54 | 57.84 | 47.94 | 11 | 9 | 11.67 |
| 10 | 47.86 | 57.48 | 23.01 | 54.43 | 67.54 | 33.87 | 11 | 8.33 | 20 |
| 11 | 45.62 | 56.77 | 40.73 | 52.61 | 61.31 | 48.37 | 11.33 | 9 | 12.33 |
| 12 | 47.29 | 78.54 | 14.19 | 55.14 | 83.78 | 22.07 | 10 | 6.67 | 32 |
| 13 | 40.58 | 50.46 | 33.24 | 42.47 | 50.04 | 31.97 | 13.33 | 11 | 17 |
| 14 | 36.67 | 58.26 | 29.13 | 31.05 | 52.83 | 25.91 | 12.67 | 9.67 | 21 |
| 15 | 50.79 | 61.76 | 46.64 | 53.38 | 64.78 | 50.25 | 10.33 | 8.33 | 11.33 |
| 16 | 60.35 | 69.30 | 63.54 | 61.57 | 69.60 | 62.14 | 8.67 | 7 | 8.67 |
| 17 | 46.60 | 67.88 | 51.30 | 43.81 | 64.26 | 45.93 | 11.67 | 8.33 | 11.33 |
| 18 | 60.45 | 70.33 | NA | 63.68 | 73.28 | NA | 8.33 | 7 | NA |
| 19 | 65.61 | 72.94 | 71.79 | 64.47 | 73.46 | 72.39 | 8 | 7 | 7 |
| 20 | 32.30 | 37.11 | 25.68 | 37.07 | 45.54 | 29.08 | 15.67 | 10.67 | 20.33 |
| 21 | 47.25 | 68.98 | 21.89 | 48.65 | 67.98 | 34.60 | 11.33 | 7 | 19.33 |
| 22 | 76.53 | 99.60 | 65.41 | 75.22 | 95.88 | 66.44 | 6 | 4.67 | 8 |
| 23 | 61.03 | 74.24 | 55.78 | 64.77 | 84.90 | 69.41 | 8 | 6 | 8 |
| 24 | 41.25 | 52.96 | 34.96 | 36.69 | 54.61 | 25.42 | 14 | 10.33 | 18 |
| 25 | 53.57 | 66.98 | 37.36 | 52.34 | 62.50 | 36.87 | 10.67 | 8.33 | 15.67 |
| 26 | 46.03 | 46.15 | 31.23 | 42.44 | 37.07 | 22.79 | 12.67 | 13.33 | 20.67 |
| 27 | 48.91 | 57.86 | 48.51 | 49.84 | 58.59 | 49.42 | 11 | 9 | 10.33 |
| 28 | 71.88 | 94.12 | 77.75 | 71.64 | 92.52 | 76.04 | 7 | 5 | 6 |
| 29 | 65.81 | 75.17 | 50.16 | 65.49 | 74.82 | 54.48 | 7.67 | 7 | 8 |

**Supplementary Figure 2 | Overview of step length and count data.**

Individual gait data for each of the 29 patients' datasets. Rounded data are presented.

| Patient | Velocity (m/s) | | | Cadence ($s^{-1}$) | | |
|---|---|---|---|---|---|---|
| | N | F | D | N | F | D |
| 1 | 65.87 | 122.03 | 30.83 | 94.93 | 117.47 | 49.33 |
| 2 | 65.80 | 130.40 | 69.87 | 89.33 | 133.07 | 95.33 |
| 3 | 110.60 | 184.33 | 99.80 | 111.97 | 154.33 | 115.73 |
| 4 | 134.07 | 234.67 | 138.80 | 124.27 | 163.10 | 125.73 |
| 5 | 87.87 | 153.40 | 37.40 | 113.67 | 154.07 | 61.83 |
| 6 | 77.20 | 112.77 | 42.57 | 106.73 | 127.57 | 75.53 |
| 7 | 78.23 | 158.73 | 62.13 | 98.43 | 137.33 | 106.17 |
| 8 | 57.37 | 118.00 | 73.53 | 151.13 | 149.30 | 177.37 |
| 9 | 95.93 | 119.67 | 86.37 | 112.33 | 125.90 | 111.20 |
| 10 | 100.83 | 158.43 | 36.53 | 117.93 | 151.60 | 76.90 |
| 11 | 110.83 | 162.27 | 73.77 | 135.57 | 164.20 | 97.10 |
| 12 | 90.00 | 164.57 | 36.97 | 105.40 | 121.97 | 123.80 |
| 13 | 65.33 | 96.30 | 41.17 | 94.23 | 114.87 | 75.53 |
| 14 | 60.80 | 119.73 | 58.33 | 107.57 | 129.27 | 127.20 |
| 15 | 92.97 | 132.93 | 85.47 | 107.00 | 125.97 | 105.57 |
| 16 | 110.07 | 144.47 | 119.10 | 108.27 | 124.77 | 113.83 |
| 17 | 79.93 | 168.07 | 82.90 | 106.33 | 152.80 | 101.77 |
| 18 | 123.93 | 173.43 | NA | 119.63 | 144.47 | NA |
| 19 | 116.00 | 154.73 | 132.27 | 107.03 | 126.87 | 110.03 |
| 20 | 55.03 | 88.67 | 42.63 | 95.40 | 129.17 | 93.33 |
| 21 | 94.90 | 143.53 | 44.40 | 118.70 | 125.53 | 89.20 |
| 22 | 133.93 | 221.07 | 117.83 | 105.90 | 135.47 | 107.00 |
| 23 | 117.37 | 183.93 | 113.47 | 111.97 | 138.67 | 106.20 |
| 24 | 68.90 | 111.30 | 55.80 | 106.40 | 123.77 | 109.90 |
| 25 | 96.30 | 158.17 | 58.10 | 109.17 | 146.87 | 93.10 |
| 26 | 74.37 | 82.50 | 49.87 | 100.90 | 119.50 | 110.87 |
| 27 | 89.00 | 133.13 | 72.87 | 108.23 | 137.20 | 89.27 |
| 28 | 125.83 | 216.27 | 134.60 | 105.17 | 139.17 | 104.97 |
| 29 | 123.77 | 204.77 | 85.67 | 113.03 | 162.83 | 98.23 |

**Supplementary Table 3 | Overview of gait velocity and cadence.**

Individual gait data for each of the 29 patients' datasets. Rounded data are presented.

| Patient | Stance % left | | | Stance % right | | |
|---|---|---|---|---|---|---|
| | N | F | D | N | F | D |
| 1 | 68.20 | 64.43 | 76.37 | 68.00 | 61.87 | 73.80 |
| 2 | 71.40 | 64.53 | 71.07 | 68.43 | 63.10 | 68.93 |
| 3 | 62.07 | 56.60 | 62.57 | 63.57 | 58.70 | 64.10 |
| 4 | 60.50 | 54.70 | 60.30 | 64.53 | 57.37 | 64.93 |
| 5 | 66.87 | 62.03 | 73.23 | 65.33 | 62.07 | 74.60 |
| 6 | 69.40 | 66.70 | 75.03 | 69.23 | 67.00 | 80.37 |
| 7 | 70.80 | 66.83 | 74.00 | 70.43 | 65.13 | 73.80 |
| 8 | 70.80 | 67.17 | 71.30 | 72.00 | 65.23 | 71.50 |
| 9 | 68.03 | 66.60 | 68.63 | 68.63 | 68.43 | 70.50 |
| 10 | 67.17 | 65.30 | 77.97 | 68.07 | 66.27 | 78.27 |
| 11 | 68.70 | 65.10 | 72.17 | 68.17 | 65.20 | 70.97 |
| 12 | 65.80 | 62.27 | 78.53 | 70.17 | 66.30 | 84.90 |
| 13 | 68.83 | 66.50 | 71.93 | 66.93 | 63.87 | 69.03 |
| 14 | 71.77 | 65.77 | 73.20 | 73.30 | 68.50 | 75.20 |
| 15 | 64.57 | 61.40 | 66.53 | 60.97 | 59.67 | 61.17 |
| 16 | 65.00 | 62.87 | 63.37 | 64.97 | 62.80 | 65.53 |
| 17 | 68.87 | 61.47 | 67.83 | 64.57 | 61.23 | 65.87 |
| 18 | 65.20 | 63.20 | NA | 63.27 | 59.20 | NA |
| 19 | 64.33 | 62.10 | 62.10 | 63.77 | 62.40 | 66.00 |
| 20 | 71.70 | 70.93 | 75.67 | 65.33 | 62.17 | 68.70 |
| 21 | 69.13 | 62.80 | 82.73 | 69.30 | 63.37 | 81.70 |
| 22 | 62.77 | 58.63 | 64.00 | 63.50 | 61.23 | 63.63 |
| 23 | 65.50 | 62.67 | 59.03 | 67.37 | 62.30 | 70.80 |
| 24 | 72.37 | 68.07 | 75.13 | 74.47 | 68.70 | 76.73 |
| 25 | 64.73 | 59.33 | 69.57 | 66.03 | 61.07 | 70.00 |
| 26 | 70.60 | 70.63 | 76.80 | 66.90 | 67.70 | 74.20 |
| 27 | 67.20 | 64.70 | 67.83 | 65.00 | 62.67 | 64.43 |
| 28 | 63.20 | 60.50 | 62.33 | 63.70 | 59.93 | 63.63 |
| 29 | 64.33 | 59.57 | 67.87 | 64.17 | 59.47 | 68.30 |

**Supplementary Table 4 | Overview of each leg's percentage of stance during the step cycle.** Individual gait data for each of the 29 patients' datasets. Rounded data are presented.
